# Environmental chemical-wide associations with immune biomarkers in the US: A cross-sectional analysis

**DOI:** 10.1101/2022.03.22.22272789

**Authors:** Lauren Y. M. Middleton, Vy K. Nguyen, John Dou, Sung Kyun Park, Justin A. Colacino, Kelly M. Bakulski

## Abstract

Exposure to environmental chemicals influence immune system functions, and humans are exposed to a wide range of chemicals, termed the chemical exposome. Thus, a comprehensive analysis of the effects across multiple chemical families with immune biomarkers is needed. In this study, we tested the associations between environmental chemicals and immune biomarkers. We analyzed the United States cross-sectional National Health and Nutrition Examination Survey (NHANES 1999-2018). Chemicals were measured in blood or urine (198 chemicals, 17 families). Immune biomarkers included percentages of lymphocytes, neutrophils, monocytes, basophils, and eosinophils, and counts of red blood cells, white blood cells, and mean corpuscular volume. We conducted survey-weighted, multivariable linear regressions of log_2_-transformed chemicals on immune measures, adjusted for age, sex, race/ethnicity, poverty-income ratio, waist circumference, cotinine concentration, creatinine for urinary chemicals, and survey cycle. We accounted for multiple comparisons using a false discovery rate (FDR). Among 45,528 adult participants, using survey weights, the mean age was 45.7 years, 51.4% were female, and 69.3% were Non-Hispanic White. There were 65 chemicals associated with white blood cell count. For example, a doubling in the concentration of blood lead was associated with a decrease of 61 white blood cells per µL (95% CI: 23–99; FDR=0.005). 122 (61.6%) chemicals were associated with at least one of the eight immune biomarkers. Chemicals in the Metals family were associated with all eight immune measures. Concentrations of a wide variety of biomarkers of exposure to chemicals such as metals and smoking-related compounds, were highly associated with immune system biomarkers, with implications for immune function and toxicology. This environmental chemical-wide association study identified chemicals from multiple families for further toxicological and epidemiological investigation.

## Introduction

The immune system is dynamic and complex, consisting of diverse cell types with varying forms and function. Proper function of immune cells in the context of a system is essential – as dysregulation has been linked to poor vaccine efficacy, increased susceptibility to infection, autoimmune diseases, and cancer.^1–4^ Clinically, immune system function is screened through measurement of blood cell populations, including quantification of lymphocytes, neutrophils, monocytes, basophils, eosinophils, red blood cells (RBCs), white blood cells (WBCs), and measurement of mean corpuscular volume (MCV – a measure of the average volume of RBCs). These immune measures are influenced by several individual-level factors including age,^5,6^ sex,^5,7,8^ and genetics.^5,9,10^ Heritability studies of the variation in these measures estimate that genetics only contribute 25-50% of the population variation.^5,9–11^ With a projected 5.08 million deaths due to top infectious diseases in 2030^12^ and the current six million (March 2022) global deaths due to COVID-19,^13^ it is critical to identify the modifiable environmental risk factors that contribute to altered immune measures.

Environmental chemical exposures can have significant impact on immune system function. Humans are exposed to environmental chemicals through air, water, food, personal care product use, industrial byproducts, or occupation. Chemicals often distribute in the blood stream, thus blood cells and the immune system become a major target for toxicological effects. For example, exposure to heavy metals, pesticides, and per- and polyfluoroalkyl substances (PFAS) are associated with immune cell proportion changes, immunosuppression, and decreased vaccine efficacy.^1,14–19^ A significant challenge in understanding how chemicals impact the immune system in humans, however, is the breadth of the possible exposures - there are more than 86,000 chemicals in commercial use in the US alone with poorly characterized immunotoxicity data.^20^ The exposome refers to considering the totality of environmental factors a person experiences with their health.^21^ While substantial progress has been made in epidemiological studies documenting associations between a few chemicals or single chemical families and immune measure outcomes,^22^ a comprehensive analysis of the effects of chemical exposures across a broad range of chemical families on immune system measures is needed.

To investigate a broad range of chemical exposures with immune biomarkers, our study tested the hypothesis that chemical exposure biomarkers would be associated with altered immune system measures, and more specifically, that chemicals from different families would have distinct patterns of immune marker dysregulation. To accomplish this, we used data from the United States cross-sectional National Health and Nutrition Examination Survey (NHANES) to test the associations between 198 chemical exposure biomarkers from 17 families of chemicals (groups of chemicals that share similar features and/or uses), with 8 immune measures across 45,528 study participants. In an exposomic analysis of the immune system, we 1) quantified associations between individual chemical exposures and specific immune measure alterations, 2) described patterns of chemical co-exposure, and 3) identified the individual chemicals and chemical families with the greatest impact on immune measures.

## Methods

### Study Participants

NHANES is a cross-sectional study conducted annually by the United States Centers for Disease Control and Prevention.^23^ NHANES is nationally representative of the non-institutionalized, civilian United States population. Data are collected by questionnaire, clinical measurements, as well as health-related biomarker measurements, including those reflecting immune system function and chemical exposures. Data on independent samples of participants are released in cycles every two years. We included ten sets of continuous cycles (1999 – 2018). Data are publicly available and supported by the National Center for Health Statistics (https://www.cdc.gov/nchs/nhanes/index.htm). Participants provided written informed consent at the time of participation. The University of Michigan Institutional Review Board (IRB) approved these secondary data analyses (HUM00116291).

### Chemical Biomarker Measures

At the time of the study visit, participants provided urine samples and trained phlebotomists collected participant venous blood in the NHANES Mobile Examination Centers. Complete laboratory methods used to measure the chemical concentrations in blood and urine and immune measures in blood are available.^24^

A total of 495 chemical biomarkers were available for initial analysis. Concentrations of all of the chemical biomarkers were not however, measured in the same cycles or in the same participants. Chemical biomarker measures were cleaned with the same process as previously established (**Figure 1**).^25,26^ Specifically, we ensured that there is one unique codename for each chemical biomarker to resolve differences in chemical codenames for the same chemical biomarker. Next, we ensured that units for concentrations were consistent over the study period for each chemical. For lipophilic chemicals measured in blood, we used the lipid-adjusted chemical measures (n=79). We excluded one measurement of urinary 4-(methylnitrosamino)-1-(3-pyridyl)-1-butanol due to no corresponding creatinine measurement for that participant.

**Figure 1.**
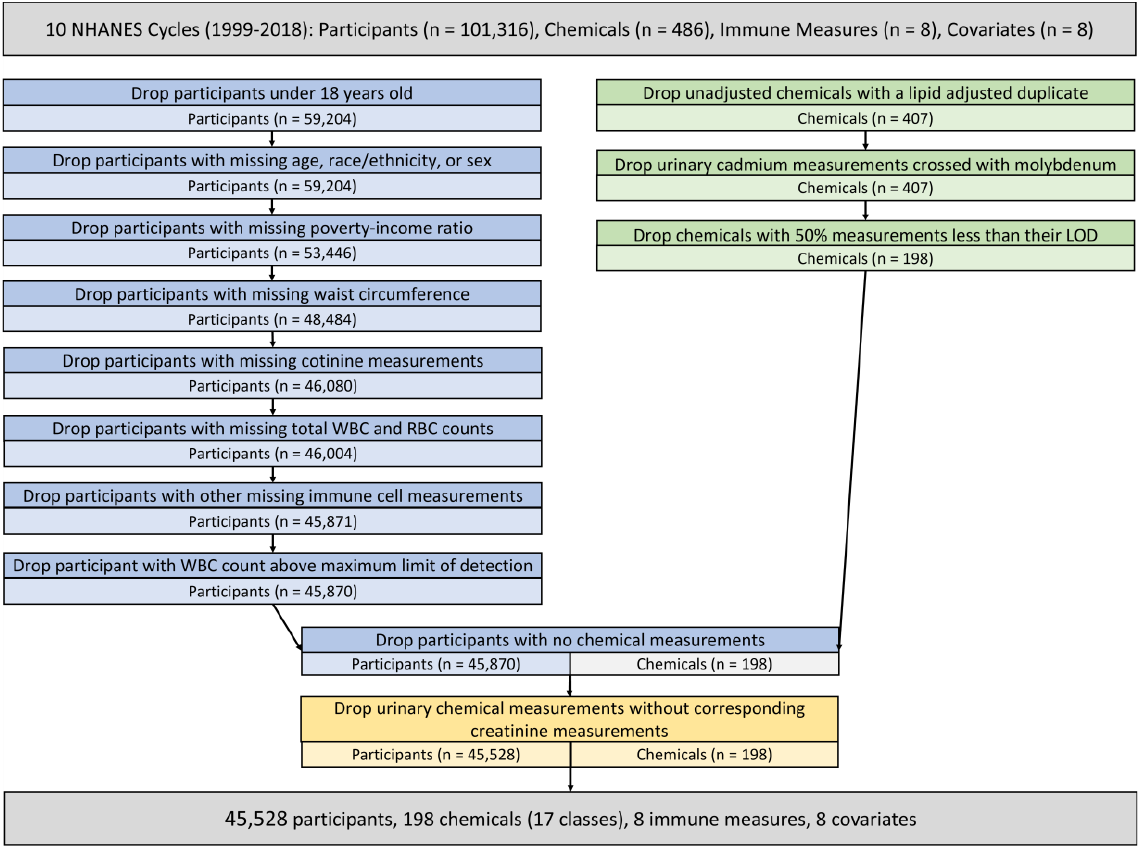
Flowchart of participant and chemical exclusion. From the 1999-2018 NHANES cycles, there were an initial 101,316 participants and 486 chemicals. The eight immune measures were percentages of basophils, eosinophils, lymphocytes, monocytes, and neutrophils and mean corpuscular volume, red blood cell (RBC) count, and white blood cell (WBC) count. The eight covariates were age, sex, race/ethnicity, poverty-income ratio, waist circumference, creatinine, smoking, and survey cycle. Participants were first excluded if they were missing covariates or immune biomarker measurements. One participant was excluded due to a WBC count above the maximum LOD. Chemicals and chemical measurements were excluded after participants. A total of 45,528 participants, 198 chemicals with 17 chemical families, eight immune measures, and eight covariates were included in this analysis.

We included chemical measures after considering the lower limit of detection (LOD) for each measure based on instrument specifications. NHANES coded measurements below the LOD as the LOD value divided by the square root of two.^27^ For chemicals with multiple comment codenames, we ensure that there is one unique comment codename per chemical biomarker. Next, we excluded chemicals with greater than 50% of their measurements below the LOD (n=209). During the study period, advances in laboratory technology influenced detection frequencies for some chemicals. To minimize such influence on our analysis, we excluded measurements that showed large changes in LOD over the study period as previously described, and used the cycles with the lowest LODs available.^25^ For example, the LODs for PCB 196 were 10.50 ng/g for cycle 2 and 0.40 ng/g for cycle 3, thus we excluded measurements from cycle 2. Our analytic sample included 196 chemicals from 17 chemical families. All chemical measures were log_2_ transformed for regression analysis. Information about each chemical, including NHANES chemical codenames, full chemical names, survey cycles in which the chemical was measured, chemical family, and limit of detection were provided in **Supplemental Table 1**.

### Immune Measures

Complete blood counts with 5-part differentials including MCV measurements were measured in the Mobile Examination Centers for all NHANES cycles with the Beckman Coulter method.^28^ The immune measures we included in this study were percentages of basophils, eosinophils, lymphocytes, monocytes, and neutrophils as well as MCV, RBC count, and WBC count. The participants included in our study sample had complete data on all immune measures.

### Demographics and Covariates

The participant covariates age (years), sex (male/female), race/ethnicity (Mexican American, Other Hispanic, Non-Hispanic White, Non-Hispanic Black, Other Race/Multiracial), and poverty-income ratio were collected by interview. We considered the following as reference groups: male, Non-Hispanic White, and the first survey cycle with measurements for each chemical. To protect anonymity, participants who reported an age greater than 85 years until 2006 and greater than 80 years from 2007-2018 were recorded in NHANES as 85 years or 80 years. We used the poverty-income ratio as a proxy for socio-economic status. The poverty-income ratio was calculated as household income divided by the poverty level for that year, adjusted for family size and inflation. Values ranged from 0 – 5 where less than 1 indicated below the poverty level, and values above 5 were rounded down to 5. Waist circumference (cm), urinary creatinine concentration (mg/dL), and cotinine concentration (ng/mL) of the participants was also collected in the Mobile Examination Centers. Creatinine and cotinine were log_2_ transformed.

### Statistical Analysis

All analyses were performed using R version 4.0.0. Code to reproduce data compilation, cleaning, and analyses are available (https://github.com/bakulskilab). To account for NHANES complex sampling design, we produced statistics representative of the non-institutionalized, civilian US population by applying survey weights to our statistical models.^26^ We selected survey weights corresponding to the smallest analysis subpopulation, and this varied by chemical measure.^29^

For the initial 101,316 participants, we compiled data on covariates (age, sex, race/ethnicity, poverty-income ratio, waist circumference, creatinine, cotinine, and survey cycle), immune system biomarkers (basophil, eosinophil, lymphocyte, monocyte, neutrophil, mean corpuscular volume, red blood cell count, and white blood cell count), and chemical biomarkers (n=486 chemicals). Participants were excluded for missing covariates and white blood cell counts above the maximum LOD. Urinary chemical measurements were excluded when missing creatinine measurements. We excluded creatinine measurements if the value was zero. We also excluded urinary cadmium measurements that were contaminated with molybdenum. We compared descriptive statistics for the demographic and immune measures on included and excluded study samples (**Supplemental Table 2**).

For each chemical, we also calculated descriptive statistics, including participant count, minimum concentration, maximum concentration, mean, standard deviation, median, interquartile range, first and 99^th^ percentiles, and percent of measurements above the LOD. To visualize the distribution of measurements above and below the LOD per chemical, we constructed a stacked bar plot. We created a scatter plot of the number of participants per chemical and percent of measurements above the LOD. To examine correlations between chemicals within and outside of chemical families, we calculated the Spearman correlations of the chemical concentrations and plotted the correlations as a heatmap. To examine correlations between immune measures, we calculated the Spearman correlations of the eight immune measures and plotted the correlations as a heatmap.

To quantify the association between chemical exposures (log_2_-transformed) and immune measure outcomes, we used survey-weighted generalized linear models that were adjusted for age (continuous), sex (categorical), race/ethnicity (categorical), poverty-income ratio (continuous), waist circumference (continuous), log_2_ urinary creatinine (continuous), log_2_ cotinine (continuous), and survey cycle (categorical) (**Equation 1**). Separate models were used for each pairwise combination of chemical and immune measure. We only included urinary creatinine in the urinary chemical models to adjust for urinary dilution.^25,30^ For chemicals that were only measured in only one survey cycle, survey cycle was not included as a covariate in the regression model. Cotinine was used as a covariate to adjust for smoking exposures.

Cotinine was also included as one of the chemical exposures, but was not used as an adjustment variable for itself. The beta coefficients from these models were reported in the text and would be interpreted as: a doubling in the chemical concentration was associated with an increase or decrease of x units of the immune measure. To account for multiple comparisons across chemicals and immune biomarkers, we calculated the false discovery rate (FDR),^31^ and we considered statistical significance as FDR<0.05. To compare the beta coefficients across chemicals with different concentration scales in the figures, we z-score standardized the log_2_-transformed chemical concentrations and conducted the same generalized linear models using the standardized, log_2_-transformed chemical concentrations. The resulting beta coefficients were used to generate the figures.

To identify patterns of association with immune measures within chemical families from the generalized linear models, we created a heatmap of the percent of chemicals significantly associated with each immune measure, separated by chemical family. We visualized the chemical beta coefficients by stratifying the results by immune measure, ordering the chemicals by chemical family, and constructing a forest plot. To visualize the strength of associations between chemical exposures and differences in immune measurements, we created a volcano plot (effect estimate versus -log_10_ FDR) for each immune measure.

Several of the chemicals in this analysis were components of cigarette smoke. As a sensitivity analysis to assess associations unadjusted for the smoking biomarker (cotinine), we ran a third set of log_2_-transformed, z-score standardized, survey-weighted, generalized linear models that were adjusted for age, sex, race/ethnicity, poverty-income ratio, waist circumference, log_2_ urinary creatinine, and survey cycle, but not cotinine. We compared the results from the cotinine adjusted models to those of the non-cotinine adjusted models in a series of Pearson correlation plots for all eight immune measures to quantify the impact of tobacco smoking on our findings.

As a sensitivity analysis, we quantified the associations between age and immune measure using a series of generalized linear models adjusted for age, sex, race/ethnicity, poverty-income ratio, waist circumference, log_2_ urinary creatinine, smoking, and survey cycle.

To understand the likelihood of a participant having a high white blood cell count compared to a normal count with exposure to blood cadmium, cotinine, or copper, we ran three separate survey-weighted logistic regressions. The chemical concentrations were log_2_-transformed, and the included covariates were age, sex, race/ethnicity, poverty-income ratio, waist circumference, log_2_ cotinine for blood cadmium and copper, and survey cycle.

## Results

### Study Participant and Chemical Descriptions

Our analytic sample size was 45,528 participants (**Figure 1**). Among participants, the survey-weighted mean age was 45.7 years (standard deviation: 17.1), 51.4% were female, and 69.3% were Non-Hispanic White (**Table 1**). The median survey-weighted values for most immune measures were within the normal adult range (**Table 2**). The percentage of monocytes, however, was slightly above the normal adult range. Compared to the included participants, excluded participants were younger (mean: 21.6 years, p<0.001), a similar percentage were female (50.8%, p=0.15), and a lower percentage were Non-Hispanic White (56.7%, p<0.001) (**Supplemental Table 2**).

**Table 1.** Survey-weighted descriptive statistics for participants (n=45,528). The participant counts were not survey weighted, and the percentages were survey weighted. The mean, standard deviation (sd), median, interquartile range (IQR), and range were calculated for the continuous variables.

|  | Count (%) | Mean (sd) | Median (IQR) | Range |
| --- | --- | --- | --- | --- |
| **Sex** |  |  |  |  |
| Female | 23,320 (51.4%) |  |  |  |
| Male | 22,208 (48.6%) |  |  |  |
| **Race/Ethnicity** |  |  |  |  |
| Non-Hispanic White | 20,283 (69.3%) |  |  |  |
| Non-Hispanic Black | 9,232 (10.4%) |  |  |  |
| Mexican American | 8,341 (8.2%) |  |  |  |
| Other Race | 4,099 (6.7%) |  |  |  |
| Other Hispanic | 3,573 (5.4%) |  |  |  |
| **Age (years)** |  | 45.7 (17.1) | 45.0 (27.0) | 18.0 - 85.0 |
| **Family Poverty-Income Ratio** |  | 3.0 (1.6) | 3.0 (3.5) | 0.0 - 5.0 |
| **Waist Circumference (cm)** |  | 98.0 (16.3) | 96.8 (21.9) | 55.5 - 179.0 |
| **Urinary Creatinine (mg/dL)** |  | 124.6 (80.8) | 111.0 (109.0) | 3.5 - 800.0 |
| **Smoking (ng/mL)** |  | 59.1 (127.0) | 0.0 (18.9) | 0.0 - 1,820.0 |

**Table 2.**
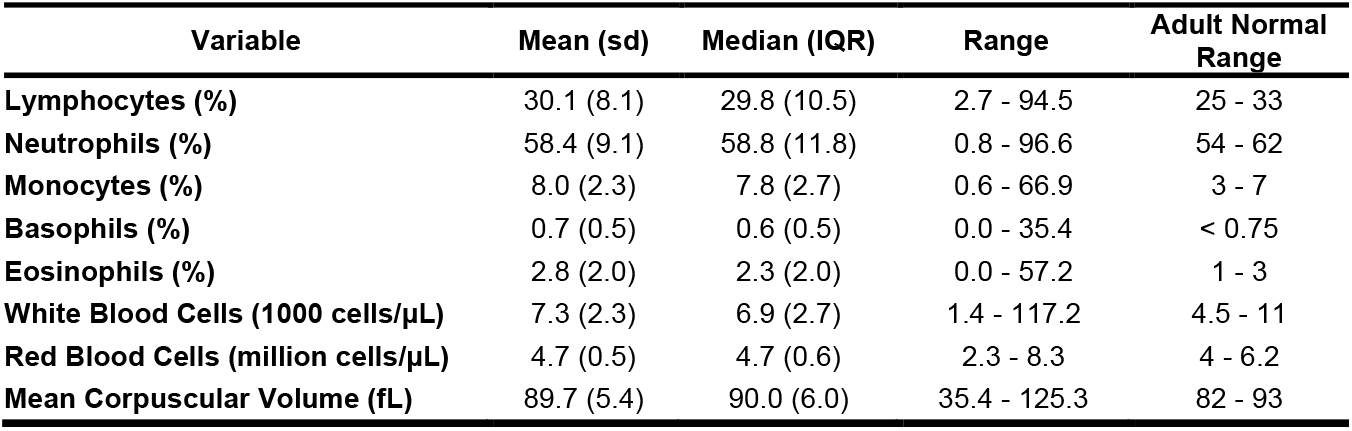
Descriptive statistics for survey-weighted immune measures (n=45,528). The mean, standard deviation (sd), median, interquartile range (IQR), and range were calculated for each immune measure. The normal range of each immune measure was reported for adults.^1^

| **Variable** | **Mean (sd)** | **Median (IQR)** | **Range** | **Adult Normal Range** |
| --- | --- | --- | --- | --- |
| **Lymphocytes (%)** | 30.1 (8.1) | 29.8 (10.5) | 2.7 - 94.5 | 25 - 33 |
| **Neutrophils (%)** | 58.4 (9.1) | 58.8 (11.8) | 0.8 - 96.6 | 54 - 62 |
| **Monocytes (%)** | 8.0 (2.3) | 7.8 (2.7) | 0.6 - 66.9 | 3 - 7 |
| **Basophils (%)** | 0.7 (0.5) | 0.6 (0.5) | 0.0 - 35.4 | < 0.75 |
| **Eosinophils (%)** | 2.8 (2.0) | 2.3 (2.0) | 0.0 - 57.2 | 1 - 3 |
| **White Blood Cells (1000 cells/μL)** | 7.3 (2.3) | 6.9 (2.7) | 1.4 - 117.2 | 4.5 - 11 |
| **Red Blood Cells (million cells/μL)** | 4.7 (0.5) | 4.7 (0.6) | 2.3 - 8.3 | 4 - 6.2 |
| **Mean Corpuscular Volume (fL)** | 89.7 (5.4) | 90.0 (6.0) | 35.4 - 125.3 | 82 - 93 |

We calculated descriptive statistics for the 198 included chemicals (**Supplemental Table 3**). Three chemicals (cotinine, blood lead, and blood cadmium,) were measured in more than 40,000 participants (**Supplemental Figure 1**). On average, smoking-related chemicals were measured in the most participants (mean=17,532) and PCBs were measured in the fewest (mean=1,365) (**Supplemental Figure 2**). Within the participants for each chemical, 16 chemicals were detected in 100% of participants and 91 chemicals were measured in ≥95% of participants.

### Chemical Concentration Correlations and Immune Measure Correlations

We calculated the Spearman correlations between the chemical concentrations (**Figure 2, Supplemental Table 4**). Smoking-related compounds (n=4) were highly correlated with themselves (mean r=0.88). PAHs (n=12) were also highly correlated with themselves (mean r=0.75). Conversely, concentrations of metals (n=26) had a low correlation with other metals (mean r=0.18). The average pairwise correlation across all chemicals was r=0.20.

**Figure 2.**
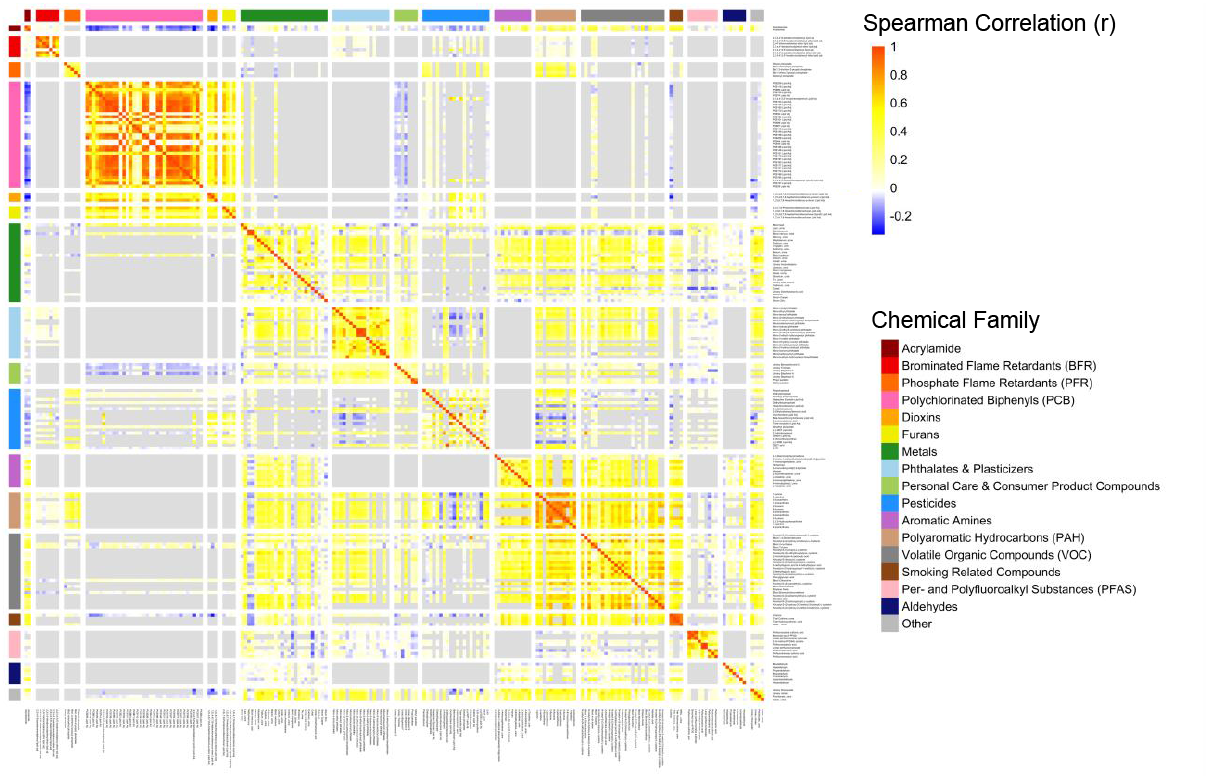
Spearman correlations between chemical concentrations. Each row and column is a chemical, grouped by chemical family as shown by the color bars. On the color scale, red indicates a more positive correlation of the chemical concentrations while blue indicates an inverse correlation. White is no correlation and grey indicates that the chemical had no participants in common and therefore no correlation could be calculated.

We also calculated the Spearman correlations between the immune measures (**Supplemental Figure 3, Supplemental Table 5**). Percentages of lymphocytes and neutrophils were highly inversely correlated (r=-0.94). The other immune measures were weakly correlated (absolute value minimum r=0.002, absolute value maximum r=0.37).

### Linear Regression Analysis

A total of 122 (61.6%) chemicals were associated (FDR<0.05) with at least one of the immune measures (**Figure 3**). All chemicals within families of acrylamides (n=2) and other (n=4) were associated with at least one immune measure (FDR<0.05). Brominated flame retardants were not associated with any immune measure (FDR<0.05). Basophils were the least likely immune measure to be associated with chemicals with only 6 (3.0%) of the chemicals associated. RBC count was associated with the highest number of chemicals with 68 (34.3%) chemicals, followed by WBC count with 65 (32.8%) (FDR<0.05). Cobalt and N-acetyl-s-(3-hydroxypropyl-1-methyl)-L-cysteine (a VOC) were both associated with all immune measures except basophils (**Figure 4, Supplemental Table 6**). Cotinine and chloroform were associated with six of the eight immune measures.

**Figure 3.**
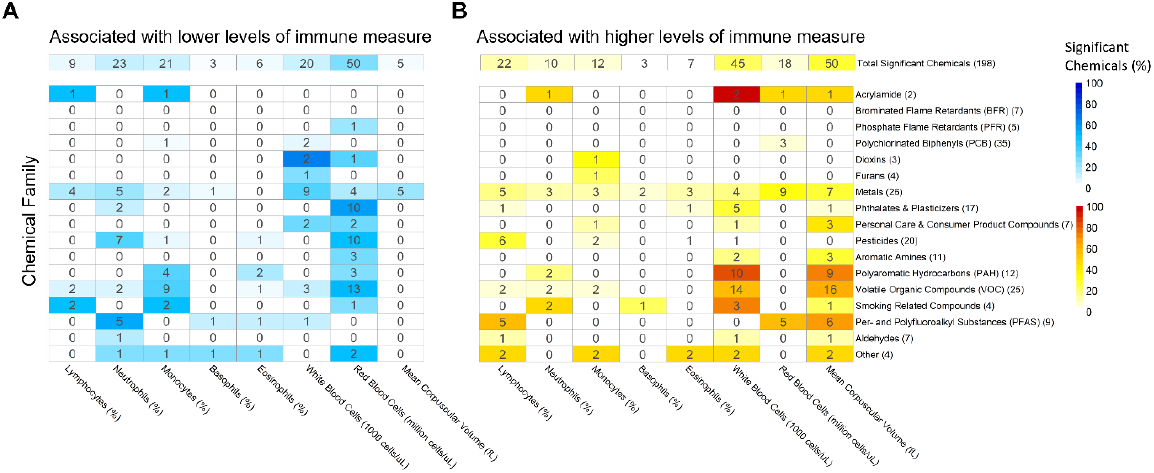
Heatmaps of significantly associated chemicals by chemical family and immune measure. The columns are the immune measures. The rows are the chemical families with the total number of chemicals per family in parenthesis in the row label. The top row is the total number of significant chemicals per immune measure. The color bars show the percent of significant chemicals out of the total chemicals in that family where dark blue or red indicates a larger percentage of significant chemicals, and white indicates no significant chemicals. The numbers in the boxes are the number of significant chemicals per immune measure and chemical family. **A)** The blue heatmap shows counts of chemicals associated with lower levels of immune measures. **B)** The red heatmap shows counts of chemicals associated with higher levels of immune measures.

**Figure 4.**
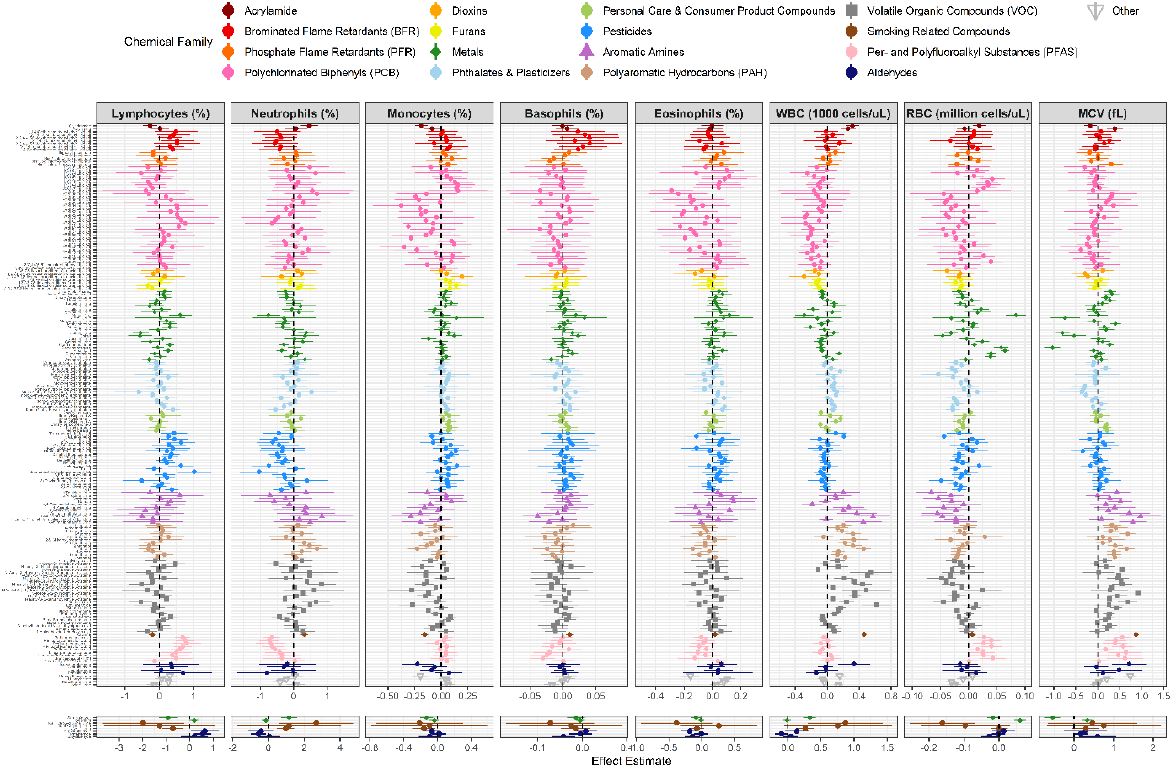
Forest plots of chemical beta coefficients from generalized linear models. Each column is a different immune measure, and each row is a chemical. The points are the beta coefficients and 95% confidence intervals. The chemicals are colored by chemical family. The separated panels at the bottom of the figure are for chemicals with effect estimates much larger than the rest of the chemicals in their column.

In the adjusted analysis, we observed 31 (25.0%) chemicals associated with lymphocyte percentage (FDR<0.05) (**Figure 3**). Of these, 22 (70.9%) of chemicals were associated with higher levels of lymphocyte percentage. Serum copper and formaldehyde were associated with the lowest and highest percentages of lymphocytes, respectively, with a doubling in the concentration of serum copper associated with 2.6% lower lymphocytes (95% CI: 1.5-3.8%; FDR=3.7×10^−4^) and a doubling in the concentration of formaldehyde associated with 2.4% higher lymphocytes (95% CI: 0.8-4.1%; FDR=0.02) (**Figure 4, Supplemental Table 6**). Chemicals in the families of acrylamides (n=1, 50.0%), metals (n=9, 34.6%), pesticides (n=6, 30.0%), smoking-related compounds (n=2, 50.0%), PFAS (n=5, 55.6%), aldehydes (n=1, 14.3%), and other (n=2, 50.05%) were associated with lymphocytes (**Figure 3**). Chemicals with the smallest FDRs were the PFAS chemicals: perfluorooctanoic acid (FDR=3.9×10^−7^), perfluorooctane sulfonic acid (FDR=3.3×10^−6^), and perfluorohexane sulfonic acid (FDR=6.9×10^−6^) (**Supplemental Figure 4**).

In the adjusted analysis, we observed 33 (16.7%) chemicals were associated with neutrophil percentage (FDR<0.05) (**Figure 3**). Of these chemicals, 10 (30.3%) were associated with higher levels of neutrophil percentage. Serum zinc and serum copper were associated with the lowest and highest percentages of neutrophils, respectively, with a doubling in the concentration of serum zinc associated with 2.9% lower neutrophils (95% CI: 1.6-4.2%; FDR=7.4×10^−4^) and a doubling in the concentration of serum copper associated with 3.3% higher neutrophils (95% CI: 2.0-4.6%; FDR=8.4×10^−5^) (**Figure 4, Supplemental Table 6**). Chemicals in the families of acrylamides (n=1, 50.0%), metals (8, 30.8%), phthalates & plasticizers (2, 11.8%), pesticides (n=7, 35.0%), PAHs (2, 16.7%), VOCs (n=4, 16.0%), smoking-related compounds (2, 50.0%), PFAS (n=5, 55.6%), aldehydes (n=1, 14.3%), and other (n=1, 25.0%) were associated with neutrophils (**Figure 3**). Chemicals with the smallest FDRs included cotinine (FDR=2.1×10^−6^), perfluorooctanoic acid (FDR=1.0×10^−5^), and perfluorooctane sulfonic acid (FDR=7.9×10^−5^) (**Supplemental Figure 4**).

In the adjusted analysis, we observed 33 (16.7%) chemicals were associated with monocyte percentages (FDR<0.05) (**Figure 3**). Of these chemicals, 12 (36.4%) were associated with higher levels of monocyte percentage. Serum copper and cobalt were associated with the lowest and highest percentages of monocytes, respectively, with a doubling in the concentration of serum copper associated with 0.4% lower monocytes (95% CI: 0.1-0.6%; FDR=0.005), and a doubling in the concentration of cobalt associated with 0.2% higher monocytes (95% CI: 0.07-0.3%; FDR=0.009) (**Figure 4, Supplemental Table 6**). Chemicals in the families of acrylamides (n=1, 50.0%), PCBs (n=1, 2.9%), metals (5, 19.2%), pesticides (1, 5.0%), PAHs (n=4, 33.3%), VOCs (9, 36.0%), smoking-related compounds (n=2, 50.0%) and other (1, 25.0%) were associated with monocytes (**Figure 3**). Chemicals with the smallest FDRs included cotinine (FDR=2.9×10^−14^), glycideamide (FDR=1.1×10^−6^), and urinary thiocyanate (FDR=5.4×10^−6^) (**Supplemental Figure 4**).

Basophils (mean=0.7%, sd=0.5%) and eosinophils (mean=2.8, sd=2.0%) were the smallest percentages of total white blood cells. There were 6 (3.0%) chemicals associated with basophil percentage (FDR<0.05). There were 13 (6.6%) chemicals associated with eosinophil percentage (FDR<0.05).

In the adjusted analysis, we observed 65 (32.8%) chemicals were associated with WBC count (FDR<0.05). Of these chemicals, 45 (69.2%) were associated with higher WBC count (**Figure 3**). Serum zinc and serum copper were associated with the lowest and highest WBC counts, respectively, with a doubling in the concentration of serum zinc associated with 1,088 fewer WBCs per µL (95% CI: 662-1,514; FDR=5.8×10^−5^) and a doubling in the concentration of serum copper associated with 952 more WBCs (95% CI: 687-1,217; FDR=1.5×10^−7^) (**Figure 4, Supplemental Table 6**). No chemicals in the BRF or PFR families were associated with WBC count. Chemicals with the smallest FDRs included cotinine (FDR=3.9×10^−64^), 2-fluorene (FDR=1.2×10^−14^), and N-acetyl-S-(2-Carboxyethyl)-L-cysteine (FDR=3.9×10^−11^) (**Supplemental Figure 4**).

In the adjusted analysis, we observed 68 (34.3%) chemicals were associated with RBC count (FDR<0.05). Of these chemicals, 18 (26.5%) were associated with higher RBC count (**Figure 3**). Cobalt and selenium were associated with the lowest and highest RBC count, respectively, with a doubling in the concentration of cobalt associated with 65,329 fewer RBCs per µL (95% CI: 43,794-86,864; FDR=3.7×10^−5^) and a doubling in the concentration of selenium associated with 321,123 more RBCs (95% CI: 229,443-412,802; FDR=1.1×10^−8^) (**Figure 4, Supplemental Table 6**). No chemicals in the BFRs, furans, and aldehydes families were associated with RBC count. Chemicals with the smallest FDRs included blood lead (FDR=1.3×10^−32^), blood cadmium (FDR=1.2×10^−17^), and blood manganese (FDR=1.7×10^−11^) (**Supplemental Figure 4**).

In the adjusted analysis, we observed 55 (27.8%) chemicals were associated with MCV (FDR<0.05). Of these chemicals, 50 (90.9%) were associated with higher MCV (**Figure 3**). Blood manganese and selenium were associated with the lowest and highest MCV, respectively, with a doubling in the concentration of blood manganese associated with 2.0fL smaller MCV (95% CI: 1.7-2.4fL; FDR=5.2×10^−14^) and a doubling in the concentration of selenium associated with 1.7fL larger MCV (95% CI: 1.0-2.5fL; FDR=4.0×10^−5^) (**Figure 4, Supplemental Table 6**). No chemicals in the BFR, PFR, PCB, dioxin, furan, and pesticide families were associated with MCV. Chemicals with the smallest FDRs included cotinine (FDR=2.6×10^−59^), urinary cobalt (FDR=1.3×10^−16^), and urinary thiocyanate (FDR=1.9×10^−14^) (**Supplemental Figure 4**).

As a sensitivity analysis to assess confounding by cigarette smoke, we compared the z-score standardized chemical beta coefficients from the regressions that included cotinine as a covariate and the regressions that were not adjusted for cotinine (**Supplemental Figure 5**). The correlations for each of the immune measure categories ranged from 0.74 to 0.98 (mean=0.90, p<1.1×10^−35^).

As a sensitivity analysis to compare the effects of a 10-year increase in age compared to increased chemical exposures, we ran generalized linear models for each of the immune measures that did not include chemical exposures as a covariate. A 10-year increase in age was associated with a 0.3 decrease in percent lymphocytes (95% CI: 0.3-0.3; FDR<1.0×10^−316^). A 10-year increase in age was associated with a 0.06 increase in percent neutrophils (95% CI: 0.05-0.07; FDR=2.2×10^−35^). A 10-year increase in age was associated with a 0.2 increase in percent monocytes (95% CI: 0.1-0.2; FDR<1.0×10^−316^). A 10-year increase in age was associated with a 0.01 increase in percent basophils (95% CI: 0.01-0.02; FDR<1.0×10^−316^). A 10-year increase in age was associated with a 0.08 increase in percent eosinophils (95% CI: 0.08-0.08; FDR<1.0×10^−316^). A 10-year increase in age was associated with associated with 12 fewer WBCs per µL (95% CI: 11-12; FDR<1.0×10^−316^). A 10-year increase in age was associated with associated with 5,408 fewer RBCs per µL (95% CI: 5,368-5,449; FDR<1.0×10^−316^). A 10-year increase in age was associated with associated with 0.07fL larger MCV (95% CI: 0.07-0.07; FDR<1.0×10^−316^).

As another sensitivity analysis, we conducted three logistic regressions with exposure to blood cadmium, cotinine, or copper. We observed that for a doubling of the concentration of blood cadmium, there was 1.16 times higher odds (95% CI: 1.09-1.24, p<0.001) of high WBC count compared to normal WBC count. For a doubling of the concentration of cotinine, there was 1.10 times higher odds (95% CI: 1.08-1.11, p<0.001) of high WBC count compared to normal WBC count. For a doubling of the concentration of copper, there was 2.58 times higher odds (95% CI: 1.50-4.43, p=0.001) of high WBC count compared to normal WBC count.

## Discussion

Chemical exposure-linked immune system dysregulation includes poor vaccine efficacy, increased susceptibility to infection, autoimmune diseases, and cancer, but few of the total chemicals that Americans are regularly exposed to have been studied.^1–4^ We tested associations between 198 chemical exposures, across 17 chemical families, with eight immune system measures using nationally representative data from 45,528 NHANES participants. We observed 122 (61.6%) chemicals were associated (FDR<0.05) with at least one immune measure. Only percent of lymphocytes and neutrophils were highly correlated (r=-0.93), which indicated that each additional immune measure provided new information. Our results showed that in the US population, exposure to environmental chemicals across 17 chemical families was associated with differences in immune measures.

MCV is a biomarker used to differentiate types of anemia based on the average volume of RBCs.^32^ We observed 55 chemicals (27.8%) associated with MCV (FDR<0.05). Prior research on MCV and environmental chemicals have noted conflicting results. In particular, associations between blood lead and MCV have varied depending on the sample size and population.^33^ A small study of car battery workers in Iran and study of pregnant women in Mexico both observed no association between blood lead and MCV,^33,34^ but another small study of pregnant women in Iran found higher blood levels were associated with lower MCV.^35^ Higher blood lead levels of lead recycling workers in Taiwan also were associated with lower MCV.^36^ These findings were supported by toxicological research where dosing rats with a mixture of heavy metals including lead decreased MCV after 90 days.^37^ In our large, generalizable US sample, we found that a doubling of blood lead concentration was not associated with MCV (FDR<0.05). Based on these observations, MCV may decrease in response to longer-term lead exposure. MCV is measured as part of complete blood panels, and values that are smaller than 80 fL or larger than 100fL can indicate diseases including iron deficiency anemia and copper deficiencies.^32,38^ Future studies may investigate the association between environmental chemicals and MCV, adjusting for intake of iron, folate, vitamin B12, and other dietary vitamins and minerals.^39,40^ Associations between MCV and environmental chemicals outside of heavy metals have been under-studied and could be a new and interesting direction for research.

A low RBC count is an indicator of anemia while a high RBC count can indicate conditions including chronically low oxygen levels or a hormonal imbalance.^41,42^ We observed 68 (34.3%) chemicals were associated with RBC count (FDR<0.05), and 73.5% of those chemicals were associated with lower RBC counts. The directions of associations in the literature were variable. In a sample of lead-exposed workers in Japan, higher blood lead was associated with lower RBC counts,^43^ while a study of pregnant women in Iran found no association between blood lead and RBC counts.^35^ Blood lead levels of lead recycling workers in Taiwan were associated with higher RBC counts.^36^ Another study using data from NHANES found that, on average, RBC counts were lowest for people with the highest PCB concentrations.^19^ The differences in study populations and lead concentrations may explain the variation in the results across these studies. Given our finding of 68 chemicals associated with RBC count, a wider array of chemicals should be tested.

WBC counts are indicative of conditions including infections and inflammation.^42^ In our study, the chemicals with the largest absolute value effect estimates for WBCs were copper (increase of 952 WBCs/µL; FDR=1.5×10^−7^) and zinc (decrease of 1,088 WBCs/µL; FDR=5.8×10^−5^) (**Figure 4**). Copper and zinc are essential minerals that are important for immune system functions and can be ingested through food sources or exposed from occupational sources.^44–46^ One prior study of 251 adolescents observed that higher copper and zinc concentrations were associated with higher WBC counts.^47^ One rat study found no association between zinc and copper deficiency and WBC count,^48^ while another found that as zinc intake increased, WBC count decreased.^49^ *In vitro* and *in* vivo studies in humans have found that zinc deficiency and high levels of zinc supplementation inhibit immune system functions.^50^ Copper deficiency and excessive levels can also result in organ system dysfunctions.^51,52^ The NHANES participants had a range of 24.7–306.6µg/dL copper (median: 113.8µg/dL) and 40.9–232.5µg/dL zinc (median: 80.4µg/L). The reference range for serum zinc was 60 – 120µg/dL and the recommended plasma copper range was 70–140µg/dL.^51^ In prior research, zinc and copper concentrations were inversely correlated,^51^ although we did not find evidence of that in our non-survey-weighted sample (r=-0.02, p=0.2). NHANES captured both deficiencies and excessive levels of copper and zinc, representing a wide range of nutritional statuses that are associated with WBC counts. Additionally, we found that increased cotinine concentrations, a biomarker of tobacco smoke, were associated with increased WBC counts (FDR=3.9×10^−64^). Following smoking cessation, prior studies observed WBC counts decreased, compared to continued smokers.^53,54^ In addition to being a biomarker of infection and inflammation, WBC counts may also be indicative of nutritional and chemical exposures.

Neutrophils are the largest component of white blood cells and function to phagocytize bacteria and cellular debris.^42,55,56^ Although in our NHANES dataset, concentrations of BFRs in blood were not shown to be associated with percentages and counts of immune cells, molecular toxicology studies showed that BFRs affect granulocyte functions.^57,58^ Smoking was previously associated with increased neutrophil counts^59^ which matches our findings that cotinine was associated with an increase in neutrophil percentage. We found that 7 (35.0%) pesticides were associated with a lower percentage of neutrophils. Previous literature on the associations between pesticide exposures and neutrophil counts have conflicting findings. One study of agricultural workers found decreased neutrophil counts associated with pesticide exposure.^60^ Another found no difference in neutrophil count in exposed and unexposed participants.^61^ Studies of agricultural workers and hospitalized patients with pesticide poisoning found an increase in neutrophil counts compared to unexposed participants.^62,63^ Differences in doses, types of pesticides, and participant populations could all contribute to the range of results. More research is needed on the effects of chemical, particularly pesticide, exposures on neutrophils.

Monocytes are important sources for inflammatory cytokines, and there are rarely disorders that result in abnormalities in only the monocytes.^64^ In our study, we observed two (50.0%) smoking-related compounds, 9 (36.0%) VOCs, and 4 (33.3%) PAHs were associated with lower percentages of monocytes. A study of cigarette smokers (exposed to smoking-related compounds, VOCs, and PAHs) found that compared to non-smokers, smoking was associated with lower monocyte counts.^65^ In contrast, two other studies observed that smoking or number of cigarettes per day was associated with an increase in monocyte counts.^59,66^ It would be of interest to consider mixtures of these chemical families rather than the association of each individual chemical to determine overall effects.

Though chemical concentrations were not always measured in the same participants, we considered the relationships between chemicals and chemical families. In our study, pairwise chemical concentrations in 6 out of 17 chemical families had an average correlation of r>0.5 (**Figure 2**). Concentrations of chemicals within these chemicals families were highly, positively correlated. Examining the average effects for these chemical families (smoking-related compounds, acrylamides, PAHs, furans, BFRs, and PCBs), identifying one indicator chemical from the family, or creating a cumulative measure of the family may be efficient methods to reduce the number of analytical variables.^22,67–69^ Conversely, concentrations of metals were less correlated (average r=0.18). The low correlation could be due to the heterogeneous sources of exposure for metals including from the natural environment, diet, and workplace. We observed in the forest plot (**Figure 4**) that the metals had a large spread and variable direction of effect on the immune measures, so the chemical family may require careful consideration when analyzing as a group. Our initial discovery analysis of pairwise chemical and immune marker associations can lead to future studies with advanced mixtures methods.

Our study had some important limitations. NHANES has a cross-sectional survey design, thus chemical and immune biomarkers were only measured in each person one time. We cannot determine temporality or longitudinal trends within each participant. We also cannot determine causality of the relationship between chemical concentrations and immune measure differences. For example, elevated blood lead could increase the RBC count through a molecular mechanism, or if lead is stored in RBCs, an increased RBC count might result in a higher blood lead concentration. In addition, NHANES does not capture long-term exposure for all chemicals. Some chemicals, like phthalates in urine, have a half-life of under an hour, while others such as cadmium in urine and lead in bones have half-lives of almost 30 years.^25^ It is possible that this study is affected by a survivorship bias. Potential participants who had the highest levels of chemical exposures may not have survived to be included in NHANES. Future studies could examine these chemicals in a longitudinal study to measure changes over time.

We excluded chemicals with less than 50% of their measurements beyond the LOD, but it is likely that some of these chemicals would be associated with the immune measures as well. A future analysis could test additional chemicals with immune measures. We used linear regressions to analyze the relationships between chemical exposures and immune measure outcomes, but there is evidence that chemicals have a non-linear association with other outcomes.^70–72^ When we examined plots of chemical concentrations and immune measures (data not shown), we observed non-linear associations for a few of the chemicals. Future analyses could test additional exposures and incorporate non-linear analyses into their methods.

We focused on immune biomarkers in our discovery study, and based on these findings, future work may consider additional immune health outcomes. For example, a future environment-wide association study could investigate the associations between chemicals and persistent infections in NHANES, expanding the work of a similar study on four PFAS chemicals.^2^ Prior research showed exposure to PCB and PFAS chemicals were associated with decreased antibodies against diphtheria and tetanus vaccines in children, potentially reducing vaccine efficacy.^1,73,74^ Our adult analysis had measures of all six PCBs and all five PFAS examined in the childhood and adolescent vaccine efficacy studies. We observed associations between six PCBs and monocytes, RBCs, and WBCs (FDR<0.05). We were not able to differentiate lymphocyte subtypes, so it is possible that B cell proportions decreased and the other subtype proportions increased more, or all subtypes increased in cell count, but these chemicals disrupted antibody production. Future studies could directly measure or estimate the proportions of lymphocyte subtypes, including B cells as well as antibody levels in response to PCB and PFAS chemical exposures. Other chemical exposures may also be associated with reduced humoral responses to vaccines and could be analyzed using the NHANES dataset.^75^

Several studies have documented associations in humans between a few chemicals or single chemical families and immune measure outcomes^40,59,76–80^, but there has not been a broad analysis of environmental chemical exposures and immune measures in the US. Our results were based on a large, diverse, and nationally representative subset of 45,528 participants from NHANES. The chemical biomarkers were rigorously quantified in urinary and blood biosamples in up to 45,528 participants per chemical with a minimum of 1,117 participants between 1999 and 2018. Our study was the first to broadly examine chemical exposures and immune measures in an exposome framework. We analyzed eight immune measures that were collected for each of these participants. From this study, we identified many chemicals for further toxicological and epidemiological studies. Out of 198 chemicals tested, 122 (61.6%) were associated (FDR<0.05) with at least one immune measure. Members of 16 out of 17 chemical families were associated with at least one immune measure. Globally in 2004, 4.9 million deaths (8.3%) and 86 million Disability-Adjusted Life Years (5.7%) were attributable to environmental chemicals, although this was an underestimate of the full global burden of disease as the study focused on 14 chemicals or groups of chemicals.^81^ Identifying chemicals that are associated with the immune system in biologically-relevant doses is an important step to understanding immune system dysregulation, setting health and safety recommendations for these chemicals, and promoting healthier environments.

## Data Availability

All data are publicly available through the National Center for Health Statistics (https://www.cdc.gov/nchs/nhanes/index.htm). All analyses were performed using R version 4.0.0. Code to reproduce data compilation, cleaning, and analyses are available upon reasonable request to the authors or on Github (https://github.com/bakulskilab).

## Acronyms

BFR: brominated flame retardant
FDR: false discovery rate
LOD: lower limit of detection
MCV: mean corpuscular volume
PAH: polyaromatic hydrocarbon
PCB: polychlorinated biphenyl
PFAS: polyfluoroalkyl substances
PFR: phosphate flame retardant
RBC: red blood cell
VOC: volatile organic compound
WBC: white blood cell

## Equation, Tables, and Figures

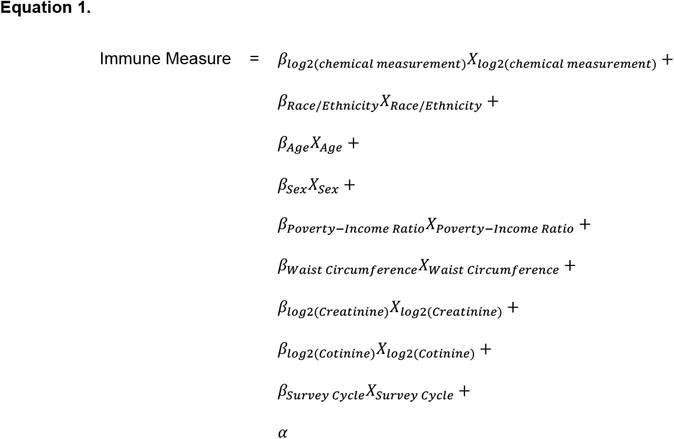

