## Supplemental Figures for "Environmental chemical-wide associations with immune biomarkers in the US: A cross-sectional analysis"

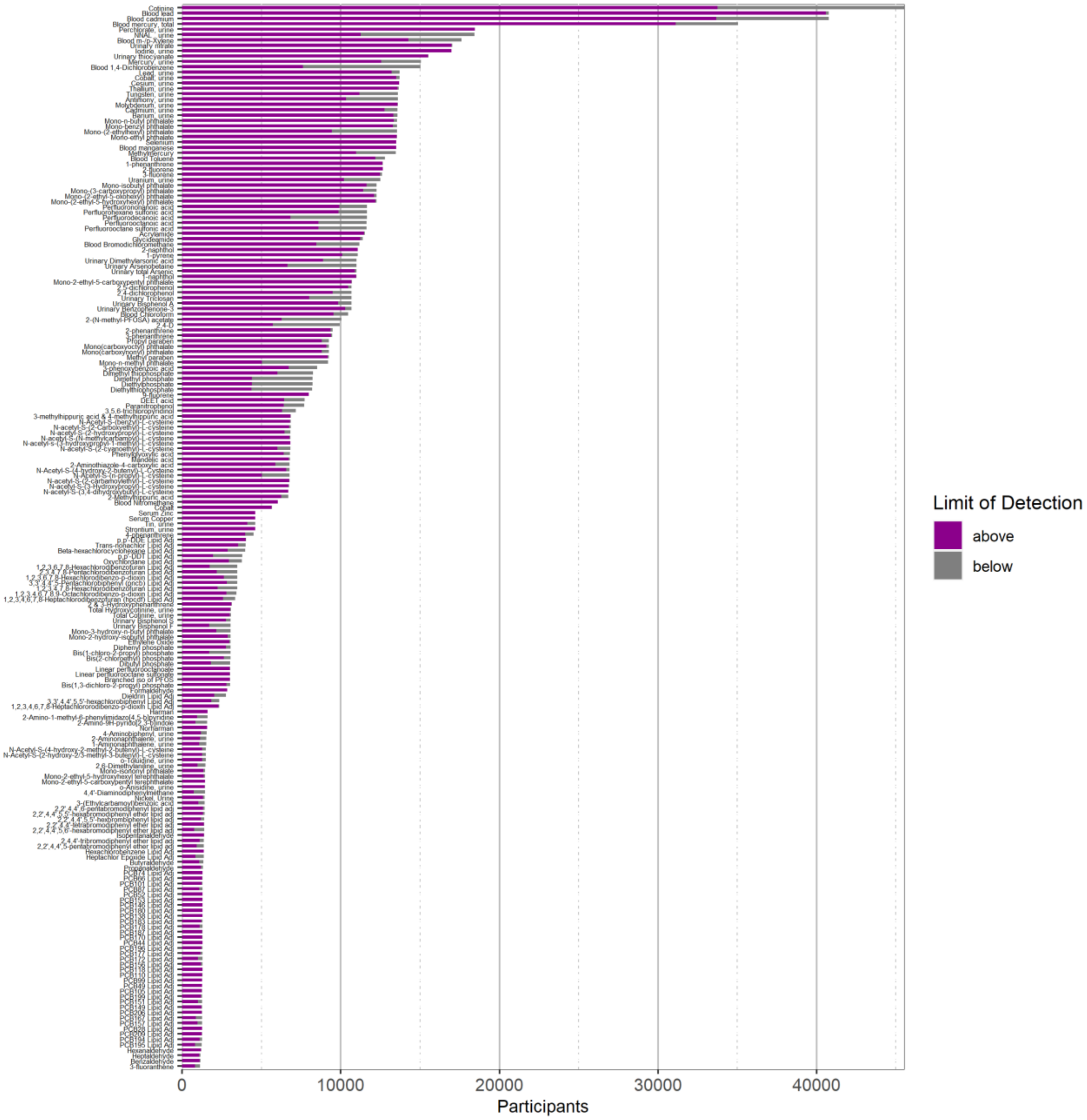


**Supplemental Figure 1.** Bar plot of number of participants per chemical, colored by number of measurements above and below the LOD for the chemicals. Purple is the number of measurements above the LOD and grey is the number of measurements below the LOD.

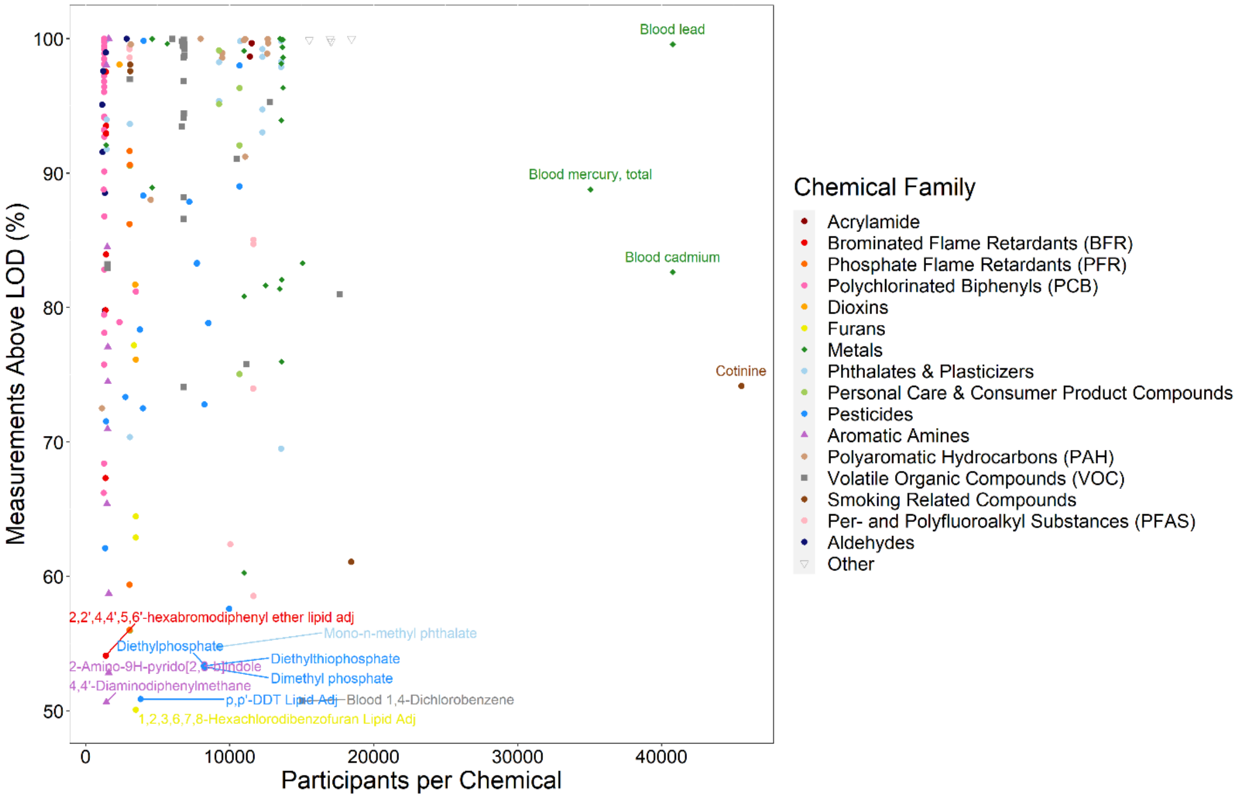


**Supplemental Figure 2.** Scatter plot of number of participants per chemical vs. percent of measurements above the LOD. Each point is a chemical, colored by chemical family. A few of the outer points are labeled with the chemical name.

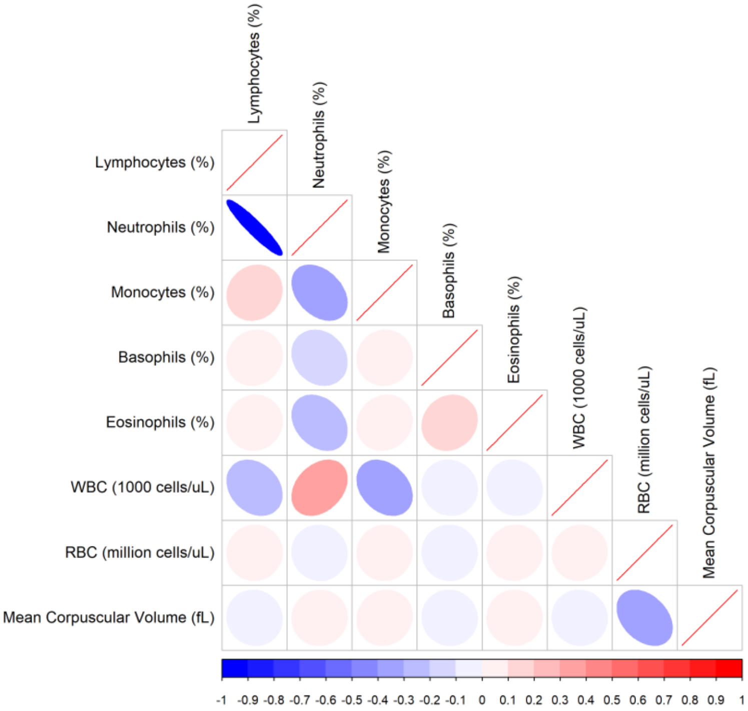


**Supplemental Figure 3.** Spearman correlation between immune measurements. The correlations are shown from blue (-1) to red (1) where values closer to the extremes are represented as more elliptical.

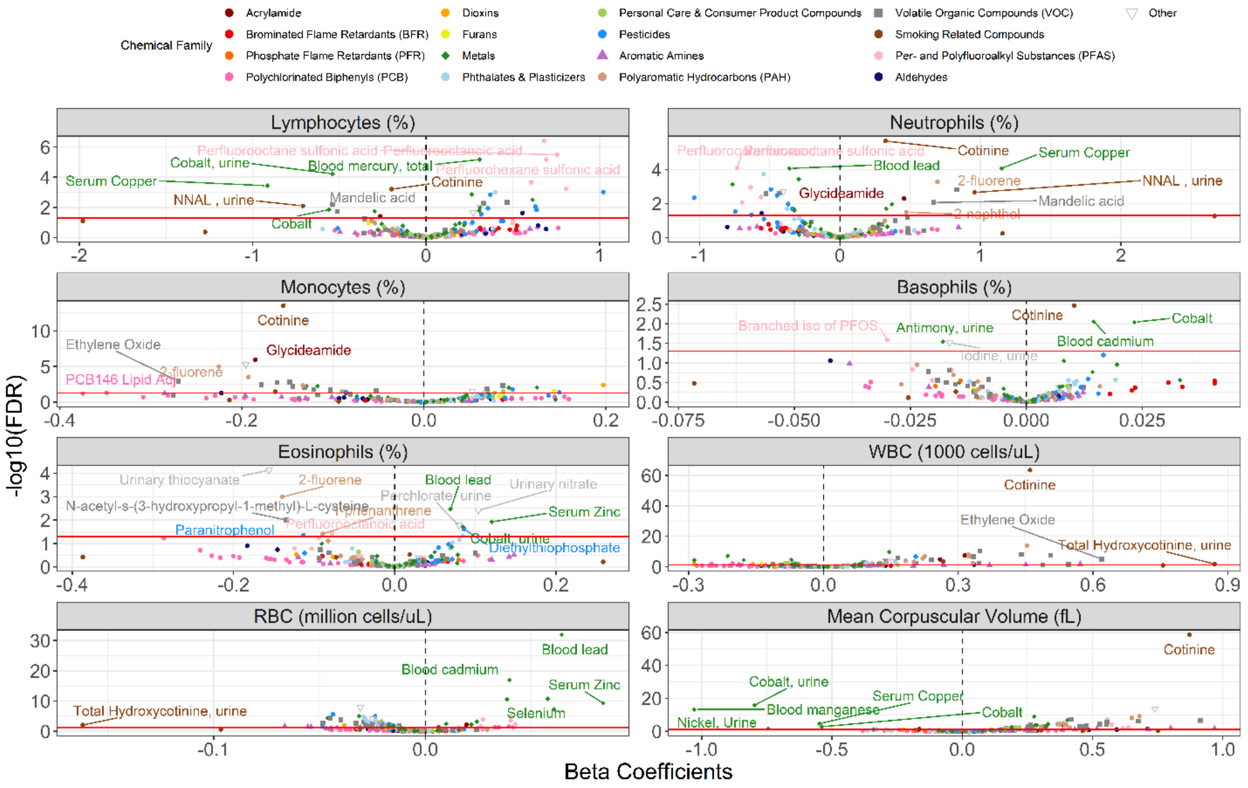


**Supplemental Figure 4.** Volcano plots of the strength of association between chemical exposures and immune measures. Each box is a different immune measure. The x-axes are the beta coefficients of the chemicals. The y-axes are the -$\log_{10}$ of the FDR for each chemical. The points are the chemicals, colored by chemical family.

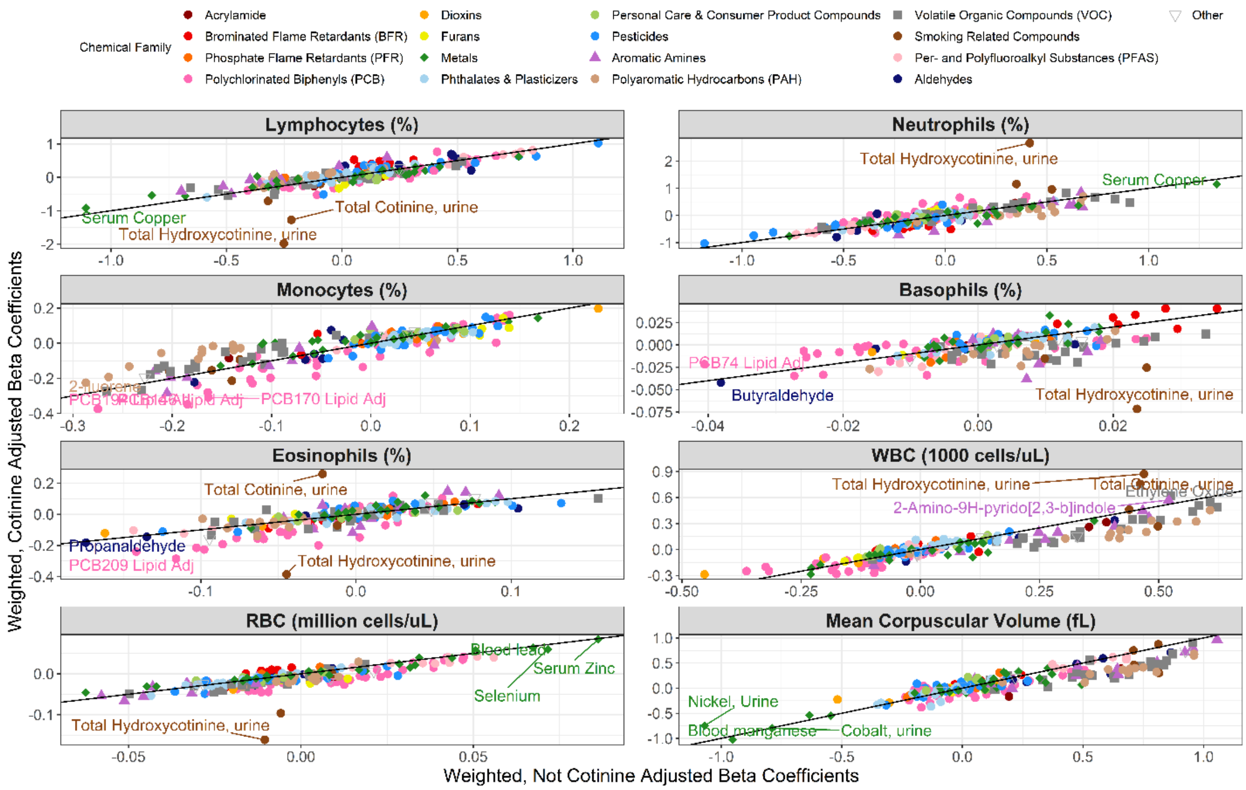


**Supplemental Figure 5.** Pearson correlation plots of beta coefficients from two sets of survey-weighted regressions adjusted for cotinine or not adjusted for cotinine. Each box is a different immune measure. The x-axis is the beta coefficients for the regressions that were not adjusted for cotinine. The y-axis is the beta coefficients for the regressions that were adjusted for cotinine. Each point is a chemical, colored by chemical family. The line represents y=x.

*Supplemental Tables*

**Supplemental Table 1.** Information about NHANES chemicals.

**Supplemental Table 2.** Descriptive statistics table of included vs. excluded study participants.

**Supplemental Table 3.** Descriptive statistics for included chemicals.

**Supplemental Table 4.** Correlations between chemical concentrations.

**Supplemental Table 5.** Correlations between immune measure values.

**Supplemental Table 6.** Results from generalized linear models.

**Supplemental Table 7.** Interpreted results from generalized linear models.
